# Prevalence And Factors Associated with Post-Caeserean Section Wound Sepsis in A Hospital in Ghana (A Retrospective Audit)

**DOI:** 10.1101/2023.10.16.23297085

**Authors:** Chukwuebuka Emmanuel, Wendy Mounibeh Bebobru

## Abstract

Caesarean Section (CS) is one of the most common obstetric procedures in Ghana. Post CS wound sepsis constitute a substantial burden to health systems in developing countries. The risk factors observed for CS wound infections are obesity, diabetes, immune-suppressive disorders, a previous CS, certain medications like steroids, the lack of pre-incision antimicrobial care, prolonged labour and surgery. Puerperal sepsis remains a notable direct cause of maternal mortality in developing and developed countries of which post CS wound sepsis contributes up to 3% especially in health units that have no facilities to carry out safe CS or treat post-CS complications. One predominant risk factor for developing puerperal sepsis is delivery by caesarean section, with some sources quoting up to a 20-fold increased risk as compared with vaginal delivery.

**Aims:** The goal of this study is to determine the prevalence and risk factors of post CS wound sepsis in Family Health Hospital.

**Methods:** This study is a retrospective audit of patient records of all women who delivered by Caesarean Section at Family Health Hospital in a 1-year period between 1^st^ January 2019 and 31^st^ December 2019. All women who delivered by caesarean section and met the inclusion criteria within the above study period were sampled. Data was collected and captured electronically by the principal researchers and then subsequently entered into Microsoft Excel and analysed using IBM SPSS version 25. The results were presented in bar charts, pie charts, frequencies and tables. Chi square test was performed on categorical data to test association between selected independent variable and the proportions of women with surgical site infection.

**Results:** The prevalence of Post-CS wound sepsis was low (5.17%) which was within the lower end of the global prevalence range (3.7% - 24.2%) and there was no statistical significance between the risk factors and post-CS wound sepsis. The number of CS done was equally distributed across the 1-year period, with an average of 9 procedures per month. Most (73%) of the women were between 31 and 40 years. The median age of the patient population was 33 years while the mean age was 33.7 years. About 63 (60%) of the women were multiparous and 43 (40%) were nulliparous at the time of CS. Only 1 (0.95%) woman was known to be HIV Positive, however there was no additional information such as when the diagnosis was made, when antiretroviral therapy was started, compliance, etc. Diabetes Mellitus was diagnosed or known in 5.71% of the women. This included both pre-gestational and gestational diabetes. The number of women who had antepartum infection was 21 which was 20 % of the study population., whilst another 20% went into labour before caesarean section was done for them. For those who went into labour, the duration of labour was not stated. Those with a history of premature rupture of membranes before CS were 7 (6.7%). Also, the number of vaginal examinations was not indicated in the clinical notes. Out of the 105 women who were identified for the study, it was difficult to find the percentage of women that had emergency CS as compared to elective CS as some of the notes lacked this.

**Conclusion:** The prevalence of post-CS wound sepsis in the hospital audited was low, and falls within the lower spectrum of the global prevalence for post-CS wound sepsis. Unfortunately, due to poor documentation, the influence of risk factors could not be clearly linked to the development of sepsis after this obstetric procedure in this hospital. Nevertheless, there is a dire need for further studies investigating post-CS wound sepsis prevalence especially in under resourced areas and tertiary centres in Ghana as there is no national data on its prevalence. This can contribute immensely in reducing maternal mortality in the nation.

## CHAPTER ONE INTRODUCTION

### 1.1 BACKGROUND OF STUDY

Caesarean Section (CS) is one of the most common obstetric procedures in Ghana. Post CS wound sepsis constitute a substantial burden to health systems in developing countries (Olsen, et al., 2009). Post CS wound sepsis is one of the most common complications for patients who have undergone CS. It is a wide spread contributing factor to maternal mortality. Some studies suggest its increase could be due to antimicrobial resistance (Muhumuza, et al., 2020). Furthermore, post CS wound sepsis can lead to septic shock if poorly managed.

The risk factors observed for CS wound infections are obesity, diabetes, immune-suppressive disorders, a previous CS, certain medications like steroids, the lack of pre-incision antimicrobial care, prolonged labour and surgery (Bhavan, et al., 2017). CS are usually done under aseptic conditions, nevertheless, the risk of post CS wound sepsis always exists which puts post CS wound sepsis as the most common nosocomial infection (Dlamini, et al., 2015). The rate of CS is increasing globally and thus, wound sepsis – the most common complication of CS is also increasing by (3-15) % (Zuarez – Easton, et al., 2017). In Ghana, the rate of CS is 6.59% and women age (30-34) years are more likely to have a CS (Manyeh et al., 2018)

In health facilities that do not have the capacity to carry out CS safely or treat post CS complications, wound sepsis is associated with maternal mortality rate of up to 3% (Mitchell, et al., 2015). In Sub-Saharan Africa post CS wound sepsis is in the percentage of 1.7% -10.4% (Chu, et al., 2012; Goldenberg, R.L., & Harrison, M.S. 2016). This has been attributed to poor access to health care services which is below 3% and thus, leading to poor postnatal follow-up and wound care (Irani, et al., 2015). In the African context, post CS wound sepsis is associated with poverty, environmental pollution, poor pre-operative care, malnutrition, anaemia, wound contamination, poor antibiotic selection and poor immunity (Gelaw, et al., 2017).

The rate of CS is below 40% in east Africa but nevertheless, this rate is high as compared to the rate by WHO which should between 10% - 15% (Worjolo, et al., 2012), therefore there is an increased risk of post CS wound sepsis in east Africa. The majority of post CS wound sepsis have pathogens that are methicillin-resistance, thus, this poses as a healthcare challenge due to the limited class of drugs available in several health centers (Kateete, et al., 2011). In addition, the lack of theatre space in many hospitals implies that theatres are being shared with other surgical teams, thus, patients with post CS wound sepsis are not isolated from patients in their wards.

It goes without saying that in order to meet the goal set by the United Nations Sustainable Development Goal, 3.1 we must aim to drastically reduce post-caesarean wound sepsis. Tackling this challenge also has benefits for the health care system since post CS wound sepsis leads to prolonged hospital stays. Implementing regular hand hygiene, keeping wounds clean and covered, and collaborating with microbiologists will mitigate the occurrence of post CS wound infection which can progress to wound sepsis. Additionally, strict aseptic preparation of surgical sites, appropriate antibiotic prophylaxis and appropriate use of theatre wear are steps in the right direction. Use of monofilament sutures, subcuticular sutures and covering surgical incisions with dressings able to absorb exudates are some of the best practices to be adopted (Krieger, et al.,2017,). Ultimately, patient educational programs are key to reducing post Cs wound sepsis by raising public awareness.

### 1.2 PROBLEM STATEMENT

Puerperal sepsis remains a notable direct cause of maternal mortality in developing and developed countries of which post CS wound sepsis contributes up to 3% especially in health units that have no facilities to carry out safe CS or treat post-CS complications (Mitchell, et al., 2015). One predominant risk factor for developing puerperal sepsis is delivery by caesarean section, with some sources quoting up to a 20-fold increased risk as compared with vaginal delivery. It leads to an increase in the average duration of stay of mothers on post-caesarean section wards, overuse of antibiotic therapy and increased hospital bills. (Ezechie et al., 2009). Currently, the prevalence of post CS wound sepsis in FHH specifically is unknown and further research is urgently needed. An unspecified number are found to have surgical site infection when they report for wound dressing and subsequently develop post CS wound sepsis. The actual rate per caesarean section in FHH is however unknown. This emphasizes the need for quantifying as well as enumerating the factors contributing to post CS wound sepsis in order to put measures in place to drastically reduce the burden on the new mothers, their newborns, families and the country as a whole (Zuarez-Easton, et al., 2017).

### 1.3 JUSTIFICATION

Post-CS wound sepsis is still a major cause of maternal mortality in the African continent and Ghana is not excluded (Manyeh, et al., 2017). In order to reduce the global maternal mortality ratio as proposed by the United Nations Sustainable Development Goal 3.1, there is a dire need to drastically reduce the numbers of women who suffer from post-CS wound sepsis (Coetzer, M., 2017). The current incidence of post-CS wound sepsis in Ghana and especially in FHH is unknown. This lacunar in our knowledge has negative consequences on maternal and child health, thus the results from this study will go a long way to influence post-CS management protocols and also make physicians more aware or conscious of the potential of post-CS wound sepsis to occur thereby, reducing the maternal and mortality in the long term (Coetzer, M., 2017).

### 1.4 AIM

The main aim of this study is to audit post caesarean wound sepsis at Family health Hospital (FHH)

### 1.5 OBJECTIVES

1. To determine the prevalence of post CS wound sepsis in FHH
2. To identify the risk factors for post CS wound sepsis in FHH

### 1.6 RESEARCH QUESTIONS

The main research question that were derived from the objectives were:

a. What is the prevalence of post-CS wound sepsis in FHH within the study period?
b. What are the risk factors for post-CS wound sepsis in FHH within the study period?

### 1.7 RESEARCH HYPOTHESIS

1. Age and Post-CS wound sepsis

H_0_: There is no statistical significance between age and Post-CS wound sepsis

H_1_: There is a statistical significance between age and Post-CS wound sepsis

2. Age and Anaemia

H_0_: There is no statistical significance between anaemia and Post-CS wound sepsis

H_1_: There is a statistic significance between anaemia and Post-CS wound sepsis

3. HIV and Post-CS wound sepsis

H_0_: There is no statistical significance between HIV and Post-CS wound sepsis

H_1_: There is a statistical significance between HIV and Post-CS wound sepsis

4. Diabetes and Post-CS wound sepsis

H_0_: There is no statistical significance between Diabetes and Post-CS wound sepsis

H_1:_ There is a statistical significance between Diabetes and Post-CS wound sepsis

5. Hypertension and Post-CS wound sepsis

H_0_: There is no statistical significance between Hypertension and Post-CS wound sepsis

H_1_: There is a significant relationship between Hypertension and Post-CS wound sepsis

6. Antepartum infection and Post-CS wound sepsis

H_0_: There is no statistical significance between Antepartum and Post-CS wound sepsis

H_1_: There is a statistical significance between Antepartum and Post-CS wound sepsis

7. Established labour prior to CS and Post-CS wound sepsis

H_0_: There is no significant relationship between Established labour prior to CS and Post-CS wound sepsis

H_1_: There is a significant relationship between Established labour prior to CS and Post-CS wound sepsis

8. Established PROM prior to labour

H_0_: There is no significant relationship between Established PROM prior to labour

H_1_: There is a significant relationship between Established PROM prior to labour

## CHAPTER TWO LITERATURE REVIEW

### 2.1 INTRODUCTION

Post CS wound sepsis is one of the most common complications for patients who have undergone CS. It is a wide spread contributing factor to maternal mortality. Some studies suggest its increase could be due to antimicrobial resistance. Furthermore, post CS wound sepsis can lead to septic shock if poorly managed (Muhumuza, et al., 2020). Given that CS rates have been increasing in developing and developed countries, it is worthwhile to consider the significant maternal and perinatal risks involved (Betran, A.P., et al., 2016)

According to the GDHS (2014), the CS rate for Ghana increased from 4.5 to 6.4 between 1990 and 2005. Also, it reported that, 13% of births were delivered by CS, an increase from 7% in 2008. Again, delivery by CS is common among women aged 35 -49 years (17%), first parity births (18%), births for women who had more than three ANC attendance (15%), deliveries in urban areas (19%), deliveries in greater Accra region (23%), birth to women with secondary school and above level of education (27%) and those with richest socio-economic status (28%) (Hafeez, et al., 2014). Women that undergoing CS are two or three times more likely to require re-admission for infectious complications than after vaginal delivery. In a bid to minimize post CS wound sepsis, common risk factors have been identified which allows for early detection of high-risk women antenally. Nevertheless, post CS endometritis and SSI are the commonest progenitors of post CS wound sepsis (Zuarez-Easton, S., 2017).

### 2.2 DEFINITIONS

**Puerperal sepsis** is defined as any temperature greater than 38 degree Celsius for more than 24hours from the day of delivery up to 10 days after (ICD 10). According to Dillen, J., et al., (2010), the clinical criteria for puerperal sepsis includes pyrexia and one of more of the following:

i. Pelvic pain
ii. Abnormal vaginal discharge
iii. Offensive discharge
iv. Delay in reduction of uterine size

The CDC (2013) defines post CS **endometritis** as temperature greater than 38 degree Celsius and/or a tender, sub involuted uterus and/or purulent/ offensive lochia and/or abdominal pain with no other recognized cause and/or diagnosis of endometritis by an attending physician.

The CDC also defines post CS **Surgical Site Infection** temperature greater than 38 degree Celsius and/or symptoms of redness, tenderness, pain and swelling around surgical site and/or purulent discharge from surgical site and/or spontaneous dehiscence of surgical wound and/or surgical wound deliberately opened by attending physician and/or abscess formation in subcutaneous tissue directly surrounding surgical wound and/or diagnosis of SSI by attending physician.

### 2.3 CURRENT GLOBAL PREVALENCE

The current post CS wound sepsis is 3.7 to 24.2% (Wloch, C., et al., 2012). Studies in some Low-and-Middle-Income Countries (LMIC) such as India and Brazil reported incidence rates of 24.2% and 11-25% respectively (Cadrso, et al.,2010). However, the post CS infection rates in High-Income-Countries (HIC) appears to be lower, for instance the National Nosocomial Infection System (NNIS) in the USA, reports an incidence of 3.5% in low risk women and 8.11% in high risk women. According to the Ghana Demographic and Health Survey (GDHS) (2014), the CS rate for Ghana increased from 4.5 to 6.4 between 1990 and 2005. Also, it reported that, 13% of births were delivered by CS, an increase from 7% in 2008. Again, delivery by CS is common among women aged 35 -49 years (17%), first parity births (18%), births for women who had more than three antenatal (ANC) attendance (15%), deliveries in urban areas (19%), deliveries in greater Accra region (23%), birth to women with secondary school and above level of education (27%) and those with richest socio-economic status (28%) (Hafeez, et al., 2014). The Saving mothers project in South Africa reports that 78% of maternal deaths due to pregnancy-related deaths are avoidable (South Africa department of Health, 2014)

### 2.4 ESTABLISHED RISK FACTORS

According to Coetzer, M., 2017, the established risk factors for post CS wound sepsis can be grouped into four categories.

1. Demographic / General Risk Factors
2. Medical Risk Factors
3. Obstetric Risk Factors
4. Surgical Risk Factors

#### 2.4.1 DEMOGRAPHIC / GENERAL RISK FACTORS

##### Body Mass Index

Increased BMI was positively correlated with an increased risk of post CS sepsis especially with SSI. A BMI greater than 35 is a significant independent risk factor for post CS SSI according to a study in England (Wloch, C., et al., 2012).

An increased subcutaneous tissue thickness greater than 2cm consequently leads to larger skin incision and more intraoperative traction, thus causing local tissue damage (Nimbalkar A.H. 2015). These factors as well as poor blood supply to adipose tissue could potentially lead to an increased risk of post CS SSI (Devjani, et al., 2013). In a retrospective study of morbidly obese women in the USA, the incidence of post CS SSI was 30% (Alanis, et al., 2010).

One study found reduced tissue concentration of cefazolin when given as pre-incisional prophylaxis in the adipose tissue of obese women. This lowered concentration poses a challenge of inadequate antimicrobial effect and highlights the need for more rigorous antibiotic cover in obese women (Pevezner, et al., 2011). A hospital-based study by Skele, E., et al (2015) demonstrated that surgical site infections occur more often among patients at nutritional risk as compared to those who are not at nutritional risk. A rigorous search of electronic data bases such as google scholar and PubMed had a paucity of data on the outcomes of underweight women post CS.

##### Younger age and Nulliparity

Younger age and nulliparity have been positively correlated with increased risk of post CS wound sepsis even though the cause remains unclear (Wloch, C., et al., 2012). This higher risk in younger women could be accounted for by lower education level, poor access to ANC, poor socio-economic status and limited support after discharge (Coetzer, M., 2017)

##### Smoking

A large retrospective study done in Washington found an increased risk of SSI in both past and current smokers (Costa, T., et al., 2018). The negative effects of smoking can be attributed to nicotine and carbon monoxide – both ultimately cause hypoxia which leads to delayed wound healing and increased risk of infections.

#### 2.4.2 MEDICAL RISK FACTORS

##### Anaemia

This is defined as Hb < 11 g/dl. According to South Africa research, anaemia was present 39.4% who succumbed to pregnancy-related wound sepsis. When severe, anaemia can cause reduced resistance to sepsis and slower recovery from infection. It is postulated that tissue hypoxia due to decreased Hb can increase anaerobic tissue metabolism thereby, increasing the potential for microbial growth (Olsen, et al., 2010).

##### Human Immunodeficiency Virus

Women living with HIV infection have higher risk of post CS wound sepsis especially when they are not yet on HAART or have low CD4 count, WHO stage 3 or 4 disease or with opportunistic infections (Coetzer, M., 2017). Given the burden of HIV in the sub-continent, prevention of MTCT and sepsis is important. Even though all HIV women are placed on HAART during pregnancy some can seroconvert and then get diagnosed at time of delivery. Ultimately, CS increases the rate of sepsis in HIV infected women (Coetzer, M., 2017)

##### Diabetes Mellitus

Diabetes is commonly associated with a higher risk of developing sepsis after a surgical procedure and post CS wound sepsis is no exception. Patients with diabetes were found to have a 2-fold increased risk of post CS SSI in Denmark (Leth, et al., 2011). A study by Schneid-Kofman in 2005 reported that pre-gestational and gestational diabetes were significant independent risk factors for post CS SSI when compared to non-diabetic women.

##### Hypertensive Disorders and pre-eclampsia

Women suffering from hypertensive disorders in pregnancy have been shown to have an increased risk of developing post CS wound sepsis (Olsen, et al., 2010). Increased capillary leakage due to endothelial disease and decreased tissue perfusion have been postulated as explanations for the relationship between pre-eclampsia and increased risk of infection (Schneid-Kofmann, et al., 2005)

#### 2.4.3 OBSTETRIC RISK FACTORS

##### Antepartum Infection

The presence of active antepartum maternal infections such as urinary tract infections and chorioamnionitis can increase the risk of post CS wound sepsis especially endometritis (Alan T.N.T. & Andrews W.W. (2010). This risk is even more significant as an incision is made into the endometrium during CS with a potential contamination of the surgical site.

##### Gestation

Preterm CS is significantly correlated with post CS wound sepsis. This is further compounded by associated factors such as pre-eclampsia (Olsen, et al.,2011) and preterm labour (Devjeni, et al., 2013).

##### Established labour prior to CS

Theoretically, this increases the risk of ascending infections from the genital tract since it is associated with cervical dilatation, vaginal examination and rupture of membranes and exposure of membranes to vaginal flora (Coetzer, M., 2017). Also, prolonged labour has been associated with post CS wound sepsis (Ghuman, et al 2011).

##### Preterm or Prolonged rupture of membranes

In a study by Devjeni, et al., (2013), 39.2% of women with prolonged rupture of membranes were found to develop post CS wound sepsis. PROM for more than 12 hours was reported to have a 2-fold risk of developing post CS wound sepsis. In a study in San Francisco, there was a significant increase in the risk of endometritis PROM for as early 12hours after rupture and not 24 hours in the previous study (Alan T.N.T. & Andrews W.W. (2010).

##### Multiple Vaginal Examinations

An Indian study found out that women with multiple vaginal examinations were not at an increased risk of SSI (Devjeni, et al., 2013). However, other studies negate this result. In Athens, a study showed an increased risk of post CS SSI in women with 6 or more vaginal examinations (Ziogos, et al., 2010). In Ohio, a study showed a significant increase in a number and type of bacterial flora as well as an increase in the amount of bacterial growth after a single vaginal examination (Alan T.N.T. and Andrews W.W., 2010). Vaginal examinations can introduce organisms into the intra-uterine structures thus, causing chorioamnionitis or post-delivery endometritis. Therefore, during incision of the uterus the organisms can spread to the incision site and cause an infection (Skele, E., et al., 2018)

#### 2.4.4 SURGICAL RISK FACTORS

##### Emergency CS

An Israeli study reported an increased risk of developing SSI when delivered by emergency CS as compared to elective CS (Scheid-Kofmann. et al., 2005). Ghuman, et al., (2011) corroborated this finding in New Zealand. Theoretically, hastened cleaning and messy surgical techniques can also lead to increased post CS wound sepsis.

##### General Anaesthesia

It has been associated with increased risk of post CS wound sepsis when compared to spinal and epidural anaesthesia (Tsai, et al., 2011). General anaesthesia is preferred in women who need emergency CS like in cases with abruptio-placentae with severe fetal distress. No large studies have been done focusing specifically on general anaesthesia and post CS infection.

##### Prolonged Surgical Time

Caesarean sections with durations exceeding 38 minutes were associated with a 2.4 times increased risk of developing post CS SSI (Opøien, et al., 2007). According to the CDC in 2013, surgical duration of more than the expected 75^th^ centile is associated with increased risk. A study in India found that, surgical duration of more than 45minutes had an SSI incidence of 53.3% (Devjeni, et al., 2013)

##### Inexperienced Surgeon

Inexperienced surgeons or surgeries done as part of training are associated with prolonged surgical time and increased risk of post CS wound sepsis (Wloch, C., et al., 2012).

##### Skin preparation

Pre-operative skin preparation with chlorhexidine in alcohol was found to be superior to iodine containing agents in preventing SSI (Tuuli, et al., 2016).

##### Cutaneous Staples

A South African study has shown a significant rise in post CS SSI due to the use of skin staples when compared to nylon or poly-glycolic sutures (Chunder, et al., 2012). This result was echoed in the USA by Figueroa, et al., in 2013.

##### Midline Incision

Midline Incisions are often performed in the emergency CS or with complicated surgeries and therefore prolonged operating time is expected (Devjeni, et al., 2013). According to the CDC in 2013, surgical duration of more than the expected 75^th^ centile is associated with increased risk.

## CHAPTER THREE METHODOLOGY

### 3.1 Study Site

This research was conducted in Family Health Hospital (FHH), a 24-hr private hospital with a 70-bed capacity located in Teshie - a coastal town in the Ledzokuku Municipal District, a district in the Greater Accra Region of south-eastern Ghana. Teshie is the ninth most populous settlement in Ghana, with a population of 171,875 people. Teshie stretches from the Kpeshie Lagoon to Teshie-Nungua Estates (first junction) from East to West on the Teshie Road. Teshie has grown enormously to become one of the biggest towns in Ghana.

The hospital functions as a general community hospital with specialist services that include obstetrics and gynecology, pediatrics, among others. The obstetric department serves a predominantly low to high income community. The staff strength of FHH is estimated as 120 health workers. It serves roughly about 156 patients annually.

### 3.2 Study Design

A retrospective audit of patient records of all women who delivered by CS at FHH in a 1-year period between 1^st^ January 2019 and 31^st^ December 2019.

### 3.3 Study Population

This research was conducted in the setting of a private community hospital in Teshie, Accra - Ghana within the study period.

#### Inclusion Criteria

All women that delivered by CS in the above 1-yearperiod will be included. Women diagnosed with post CS sepsis within 30 days after surgery was identified and records audited. The CDC criteria for Surgical Site Infection (both superficial and deep) as well as endometritis was used for diagnosis. The entire study population was used for this study because this is the first of such study and will help to accurately estimate the prevalence of post CS sepsis. The criteria are as follows:

The CDC (2013) defines post CS **endometritis** as

- temperature greater than 38 degree Celsius And one or more of the following
- a tender, sub involuted uterus
- purulent/ offensive lochia
- abdominal pain with no other recognized cause
- diagnosis of endometritis by an attending physician.

#### The CDC also defines post CS **Surgical Site Infection**

- temperature greater than 38 degree Celsius And one or more of the following
- symptoms of redness, tenderness, pain and swelling around surgical site
- purulent discharge from surgical site
- spontaneous dehiscence of surgical wound
- surgical wound deliberately opened by attending physician
- abscess formation in subcutaneous tissue directly surrounding surgical wound
- diagnosis of Surgical Site Infection by attending physician.

#### Exclusion Criteria

Women who didn’t have deliveries during the study period would be excluded as well as women whose adequate medical records cannot be retrieved.

### 3.4 Sample Method and Sample Size

Total population sampling was done. Every woman who delivered during the 1-year study period and met the inclusion criteria will be included.

Using Taro Yamane’s formula, the Sample size was calculated as follows;

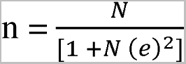

where;

n= sample size

N=population size

e = sampling error

the total number of women delivered by CS is 156 which represents the study population. Using a sampling error of 0.05

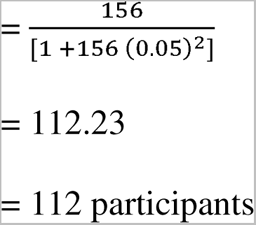

The calculated sample size for the study is 112.

Participants was identified by the principal researcher using the CS theatre register as well as cross checking in the birth register. Patients records are currently stored electronically in the hospital. All patients’ files were accessed electronically in order to obtain clinical records. Women diagnosed with post CS wound sepsis during inpatient or at follow-up was reviewed and data captured in four main categories:

- General / Demographic (age / smoking / body mass index etc.)
- Medical (Underlying medical conditions, antibiotic use etc.)
- Obstetric (Gravidity / Parity / Time from rupture of membranes / Previous CS etc.)
- Surgical (Surgical time / Surgeon experience / Indication etc.)

Patients records was manually accessed by the principal researcher in order to identify women diagnosed with post CS endometritis or surgical site infection. Patients’ information on clinical course, outcome, subsequent hospital admission, repeat surgery, severe morbidity and mortality was captured.

### 3.5 Data Management and Statistics

Data was collected and captured electronically by the principal researcher under the supervision of the study supervisor and then subsequently entered into Microsoft Excel ® and analysed using IBM SPSS version 25. The results were presented in summary tables and charts and analyzed using frequencies, means, proportions and percentages. Chi square test was performed on categorical data to test association between selected independent variable and the proportions of women with surgical site infection.

### 3.6 Privacy and Confidentiality

Patients information was kept confidentially by using a computer that is password protected. Also, data was extracted using patient’s hospital identity numbers and not their names.

### 3.7 Ethical Considerations

Ethical clearance was obtained from the University of Ghana Medical School Community Health Department Dissertations Review Committee.

## CHAPTER FOUR RESULTS

### 4.1 Introduction

This chapter presents the socio-demographic, medical, obstetric and surgical characteristics of the participants, the prevalence of post -caesarean section wound sepsis in FHH, the factors associated between the various independent variables and the outcome variable, post-caesarean section wound sepsis.A total of one hundred and fourteen (114) women were identified as having delivered by caesarean section (CS) in the study period (from 1 January 2019 – 31 April 2019). Nine (9) women were excluded, as minimal or no clinical notes were available. Of the 105 records audited, the number of CS done was equally distributed across the 1-year period, with an average of 9 procedures per month.

### 4.2 Socio-demographic characteristics of participants

The background characteristics of all the 105 women who had undergone caesarean section at FHH during the study period is as follows;

Most (73%%) of the women were between 31 and 40 years. The median age of the patient population was 33 years while the mean age was 33.7 years.

**Figure 1:**
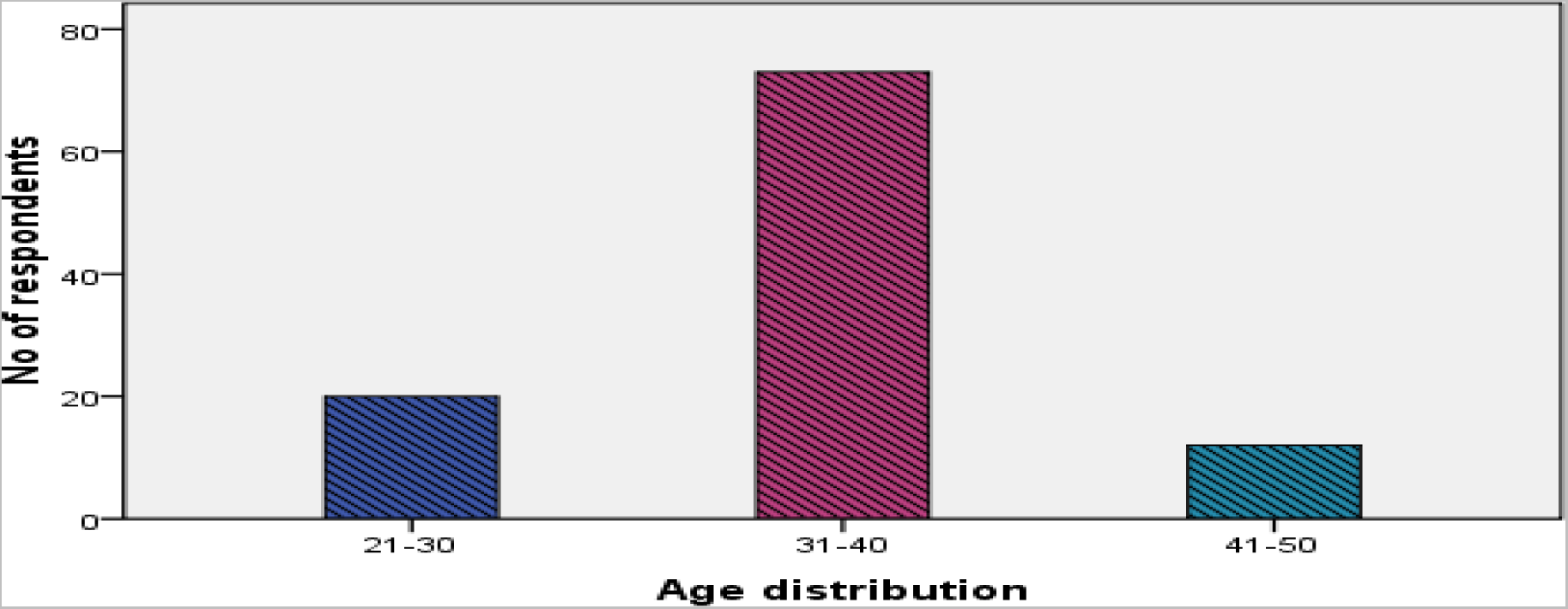
Distribution of age in the study population.

About 63 (60%) of the women were multiparous and 43 (40%) were nulliparous at the time of CS.

**Figure 2:**
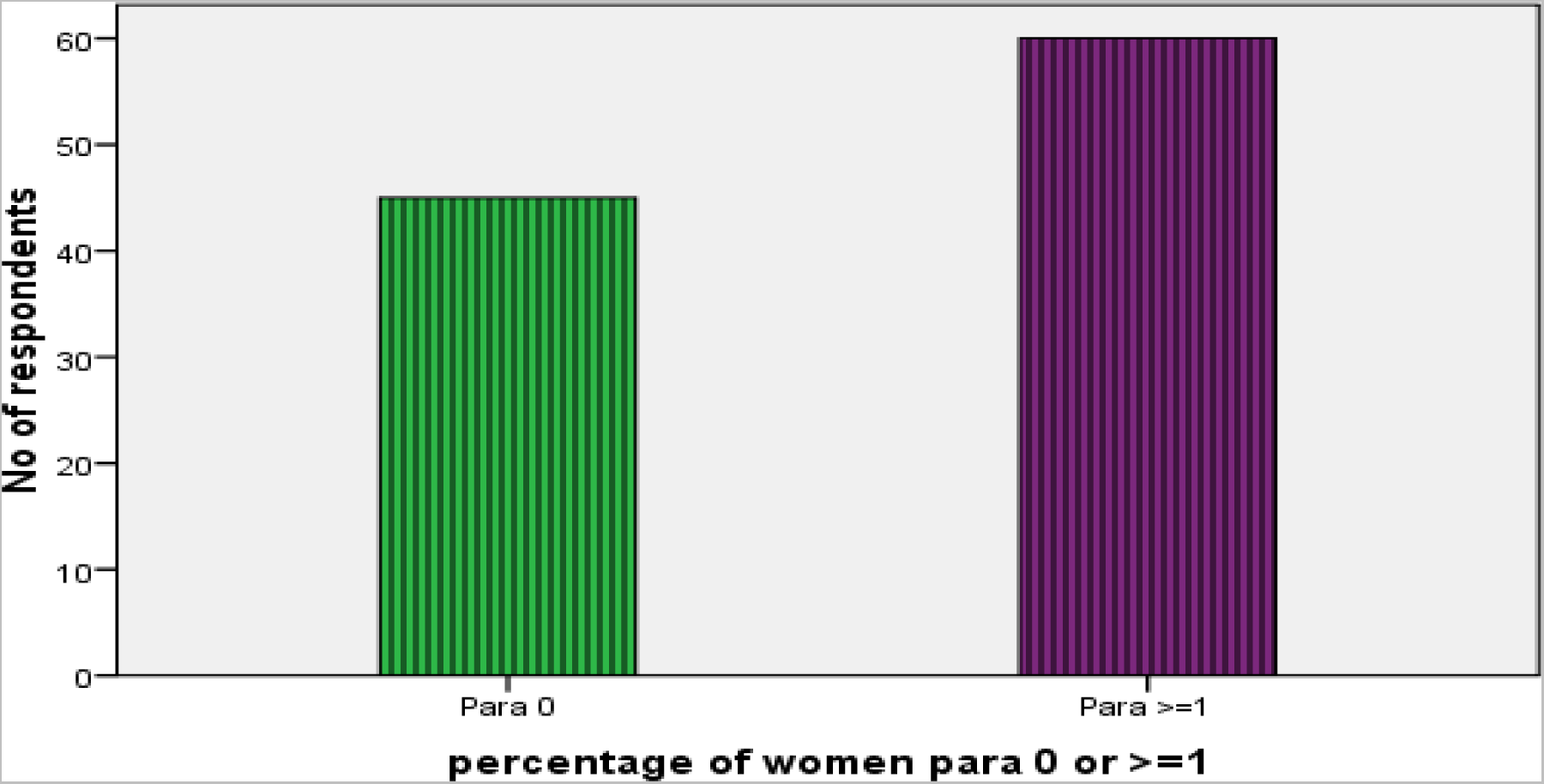
The distribution of parity in the study population.

None of the women during the period smoked cigarettes either before or during the pregnancy.

Approximately 86.7 % of the women had only their weight inputted without their respective heights and therefore BMI could not be calculated.

### 4.3 Medical Data

Majority (69.5%) of the women were not anemic (Hb > 11g/dl) before the procedure.

**Figure 3:**
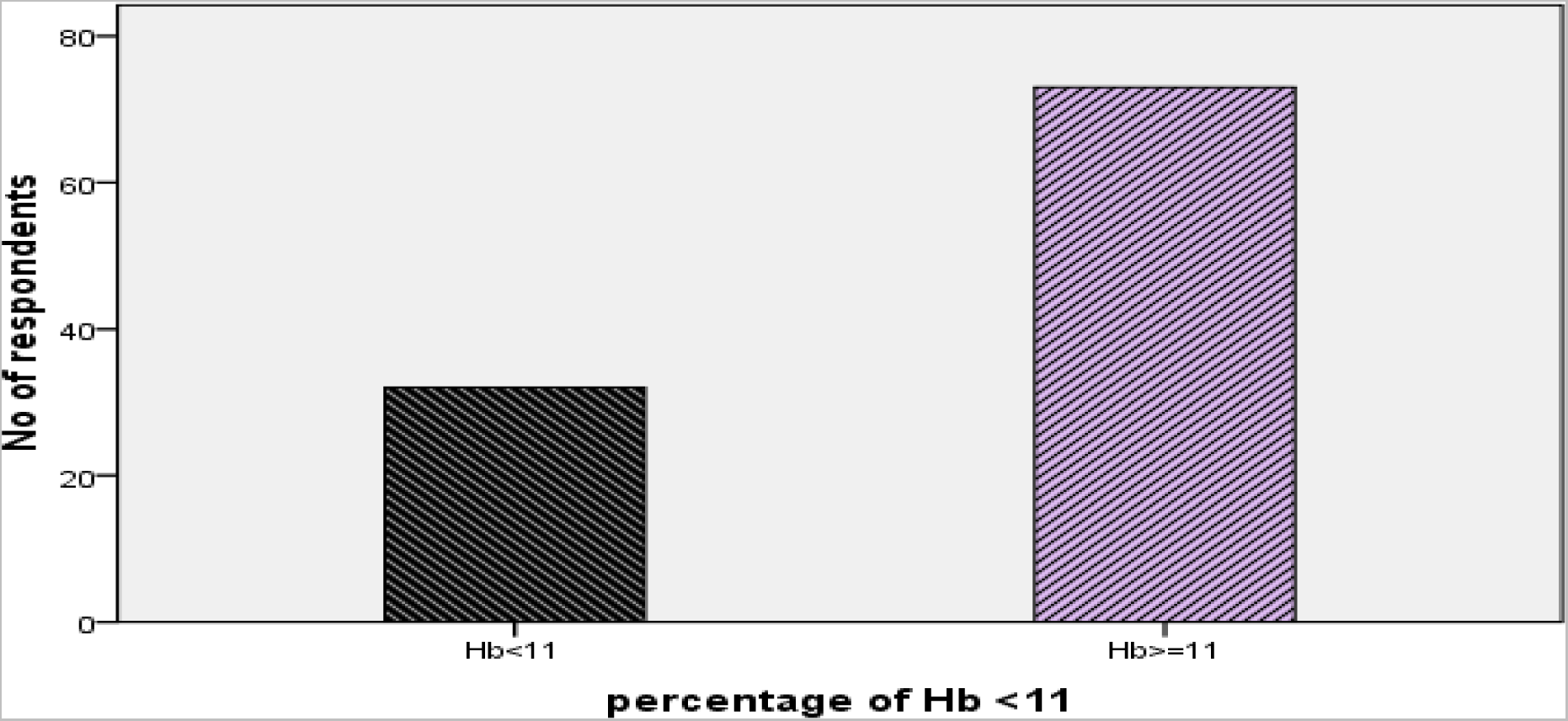
The distribution of anaemia in the group

Only 1 (0.95%) woman was known to be HIV positive, however there wasn’t any additional information such as when the diagnosis was made, when antiretroviral therapy was started, compliance, etc.

**Figure 4:**
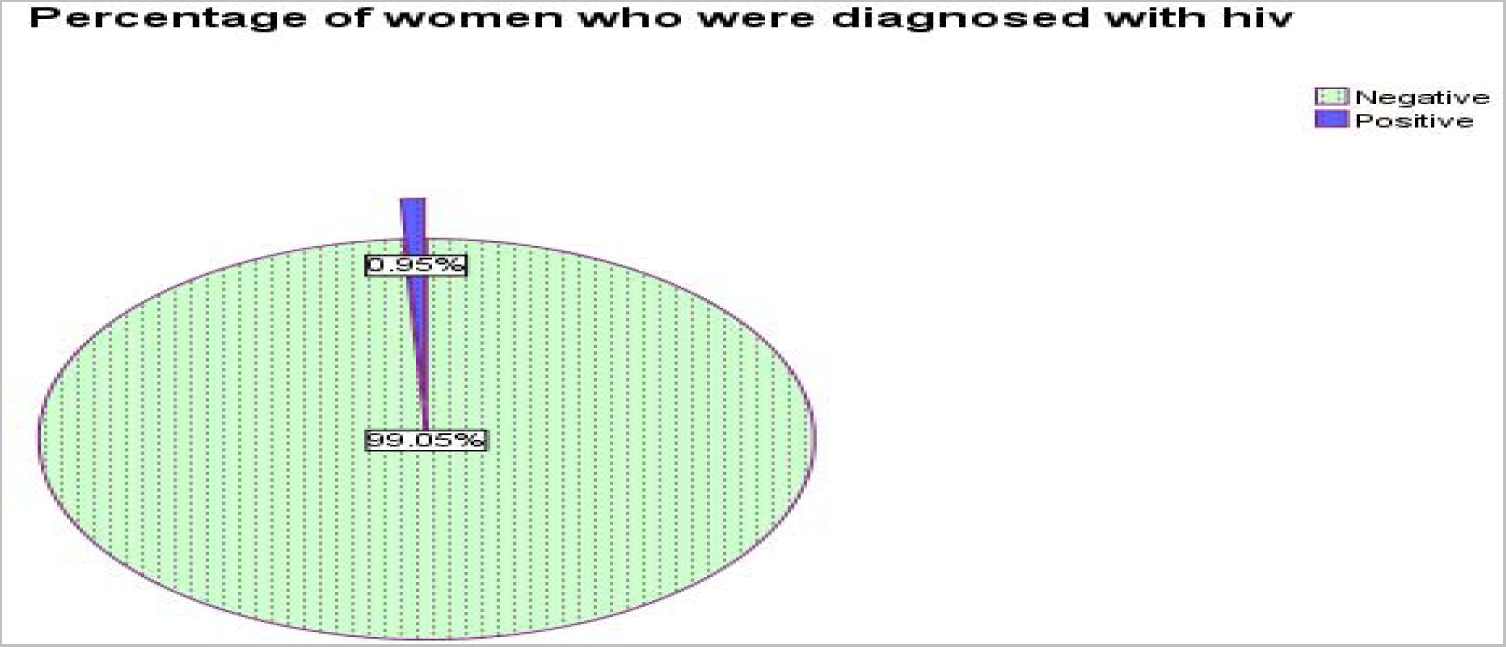
The distribution of women diagnosed with HIV in the group

Diabetes Mellitus was diagnosed or known in 5.71% of women. This includes both pre-gestational and gestational diabetes.

**Figure 5:**
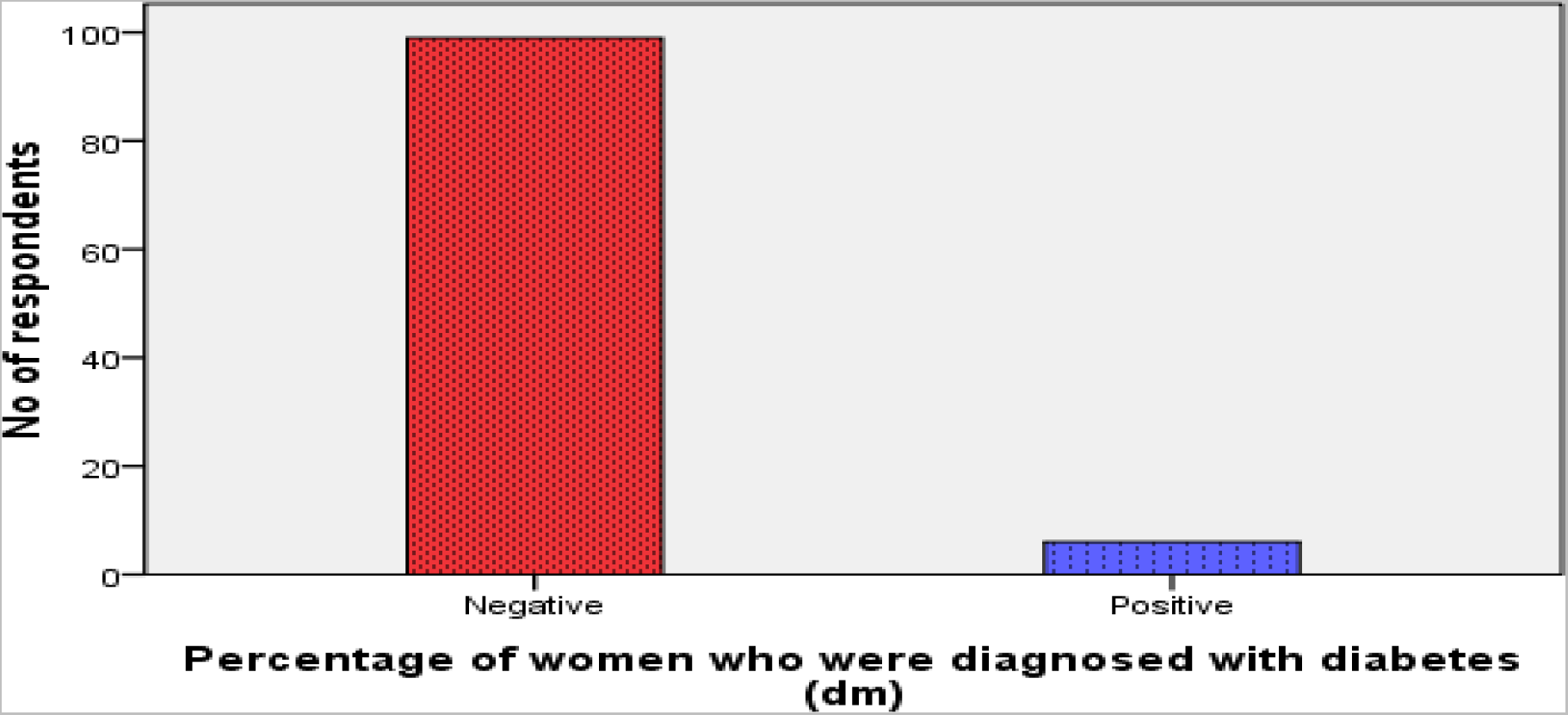
The distribution of Diabetes in the study population

Hypertensive disorders were present in 16.19% of women included, with preeclampsia mentioned in some of the notes.

**Figure 6:**
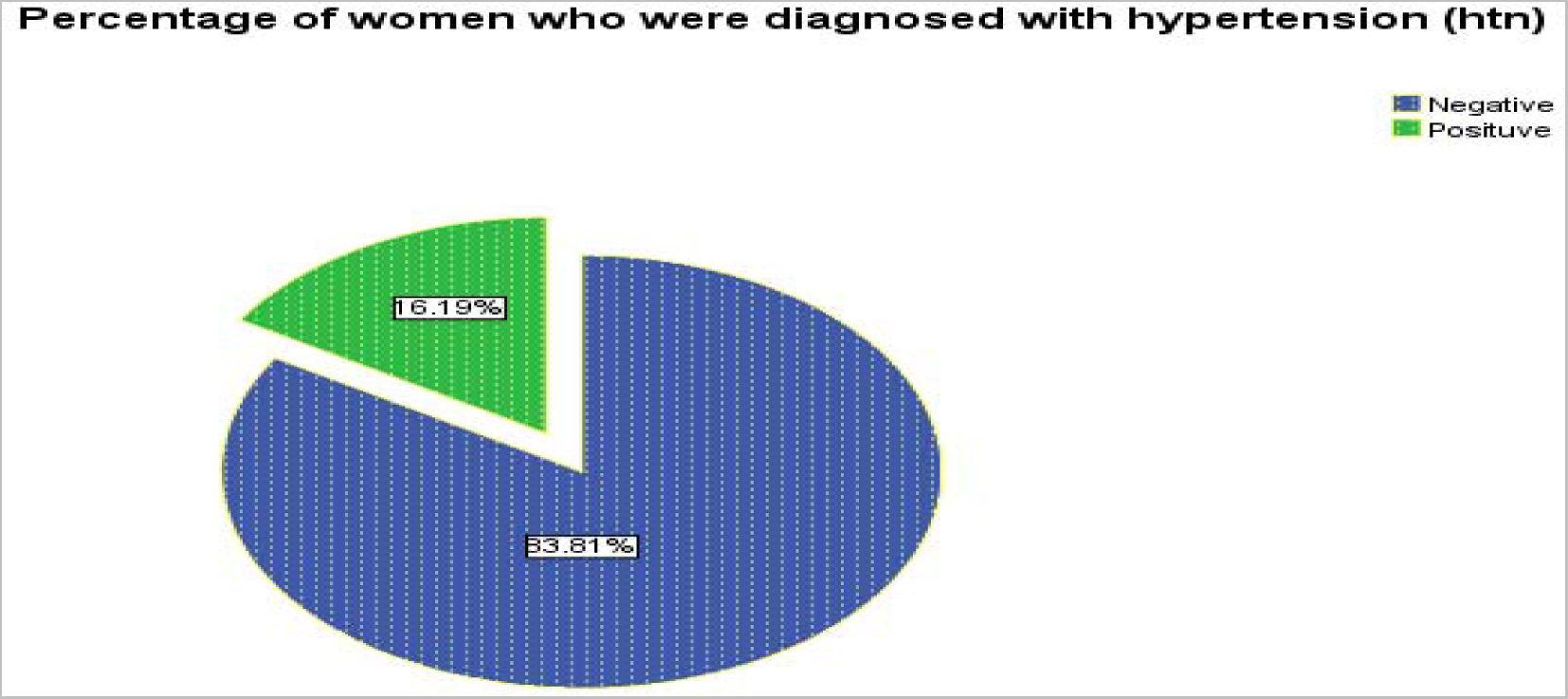
The distribution of hypertension in the group

### 4.4 Obstetric Data

The number of women who had antepartum infection was 21 which is 20% of the study population. The median gestational age (in completed weeks) at delivery could not be calculated because it wasn’t recorded for all the women but generally, it ranged from 35 to 42 weeks.

Out of the total study participants, 21 (20%) went into labour before caesarean section was done for them. For those who went into labour, the duration of the labour wasn’t stated.

**Figure 7:**
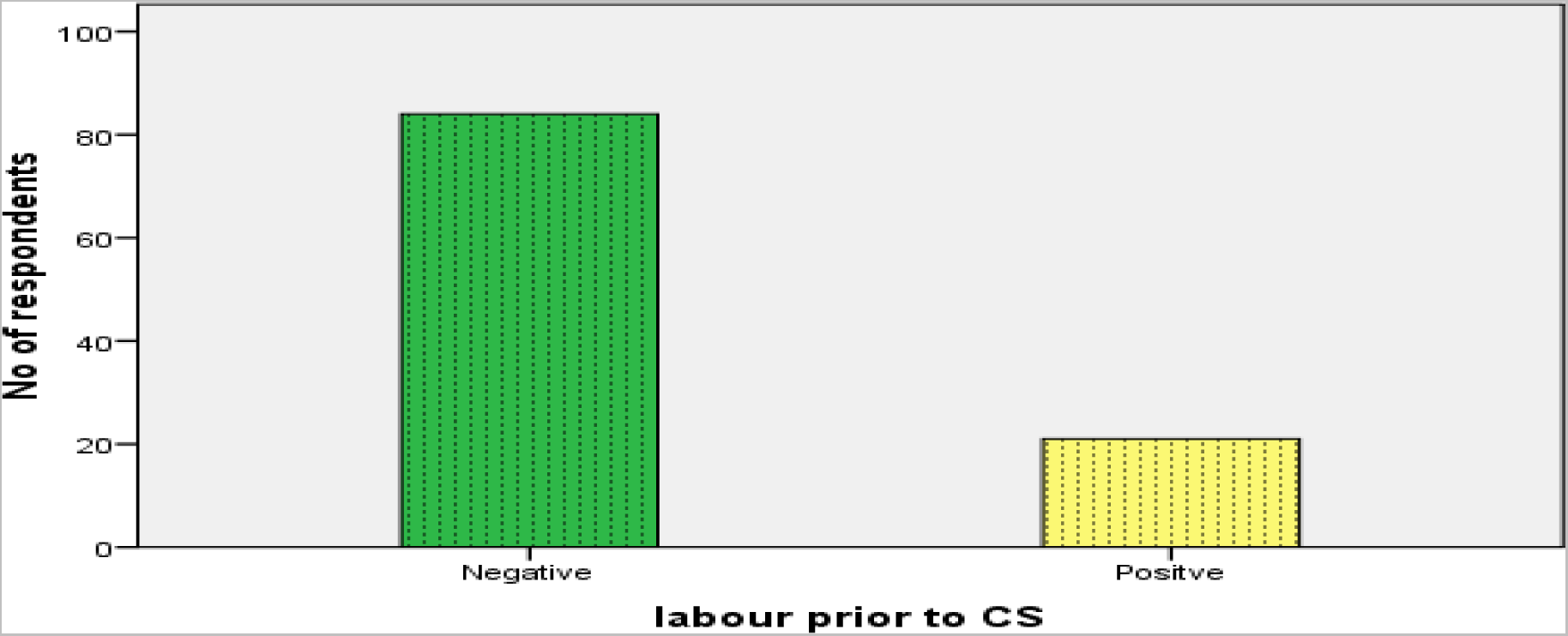
The distribution of presence of established labour prior to the procedure in the group

Those with a history of premature rupture of membranes before CS (PROM) were 7(6.7%). Also, the number of vaginal examinations wasn’t indicated in the clinical notes.

### 4.5 Surgical Data

Out of the 105 women who were identified for the study, it was difficult to find the percentage of the women that had emergency CS as compared to elective CS as some of the notes lacked this information.

The Pfannenstiel skin incision technique was used for the all the Caesarean Sections. None of the women had the midline incision type. All the procedures were done by experienced surgeons (residents and above); chlorhexidine was used for the pre-operation skin preparation and anaesthesia was spinal.

The skin closure technique and the duration of the caesarean section wasn’t indicated in the clinical notes but staples weren’t mentioned at all.

### 4.6 Prevalence of Post – Caesarean Section Wound Sepsis

During the 1-year study period, 6 patients were identified with post CS wound sepsis, which translates to a prevalence rate of 5.71% (6 out of 105).

**Figure 9:**
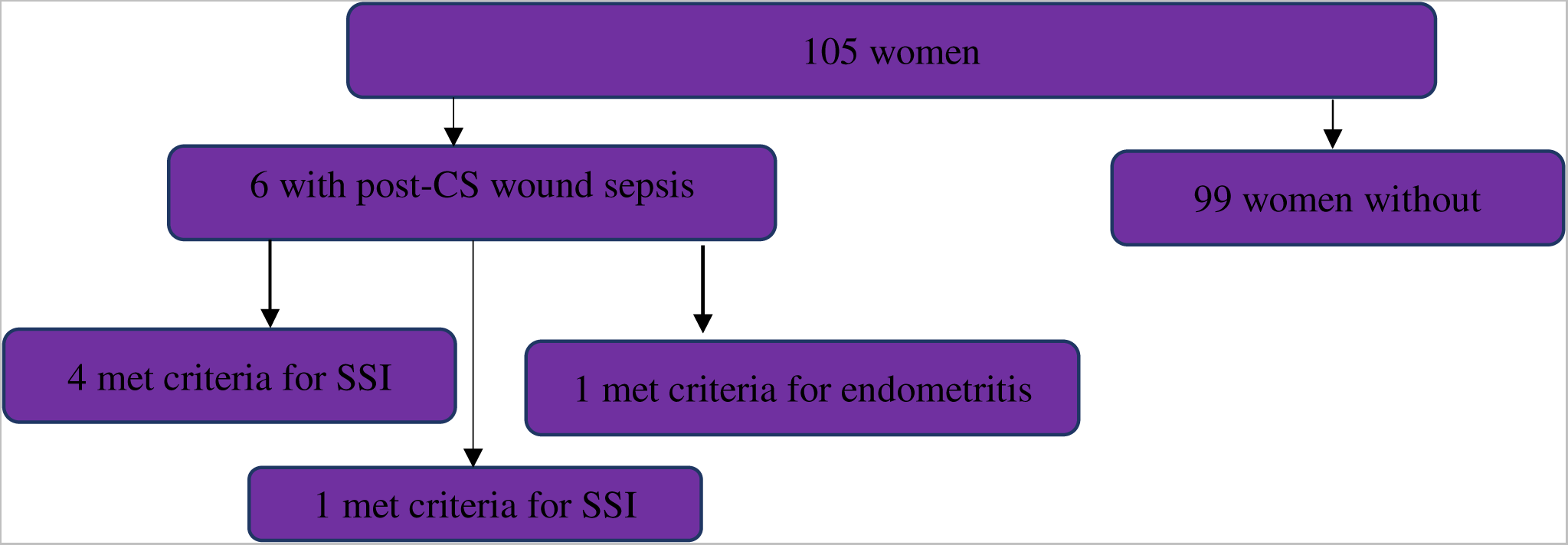
The distribution of post-CS sepsis among the study population

### 4.7 Risk factor analysis

When identifying risk factors for post CS sepsis, all patient- and caesarean section factors mentioned above were analysed as possible risk factors. All statistical tests were carried out at 5% level of significance and the alpha (α) value of 0.05. The majority of factors were found to not reach statistical significance.

**Table 1:** Age as a risk factor for post-CS wound sepsis.

| Independent variable | Post-CS wound Sepsis | | $\chi^2$<br>(Pearson,<br>Likelihood ratio) | p-value |
| --- | --- | --- | --- | --- |
| Age (Years) | yes | no | 1.886, 3.220 | 0.170, 0.383 |
| $\leq 30$ | 0 | 24 | | |
| $>30$ | 6 | 75 | | |

From the table above, the non-parametric Pearson chi-square statistic is 1.886 and the likelihood ratio statistics is 3.220 with a corresponding asymptotic p-value of 0.170 and 0.383 respectively. Nevertheless, the result shows that there is a statistically insignificant relationship between post CS wound sepsis and age distribution of women this is confirmed by their p-values of 0.170 and 0.383 respectively which is more than the alpha value of 0.05. Therefore, we fail to reject the null hypothesis that states that there is no significant relationship between post CS wound sepsis and age distribution of women and we thus reject the alternate hypothesis that there is a significant relationship.

**Table 2:** Anaemia as a risk factor for post-CS wound sepsis.

| Independent variable | Post-CS wound Sepsis | | $\chi^2$<br>(Pearson,<br>Likelihood ratio) | p-value |
| --- | --- | --- | --- | --- |
| Haemoglobin (Hb) | yes | no | 1.145, 1.058 | 0.285, 0.304 |
| Hb ≤ 11 | 3 | 3 | | |
| Hb $>$ 11 | 29 | 70 | | |

From the result above the non-parametric Pearson chi-square statistic and the likelihood ratio statistics have a corresponding asymptotic p-values of 1.145 and 1.058 respectively. However, the result shows that there is a statistically insignificant relationship between post CS wound sepsis and Hb among women this is confirmed by their p-values of 0.285 and 0.304 respectively which is less than the alpha value of 0.05. Therefore, we cannot reject the null hypothesis that states that there is no significant relationship between post CS wound sepsis and Hb among women and we reject the alternate hypothesis that there is a significant relationship. In summary therefore we can state that there is no evidence that Hb is a risk factors that may determine post CS wound sepsis among women.

**Table 3:** HIV as a risk factor for post-CS wound sepsis.

| Independent variable | Post-CS wound Sepsis | | $\chi^2$<br>(Pearson,<br>Likelihood ratio) | p-value |
| --- | --- | --- | --- | --- |
| HIV | yes | no | 0.061, 0.118 | 0.805, 0.731 |
| Yes | 0 | 6 |  |  |
| No | 1 | 98 |  |  |

From the result above the non-parametric Pearson chi-square statistic is 0.061 and the likelihood ratio statistics is 0.118 corresponding asymptotic p-values of 0.805 and 0.731respectively. However, the result shows that there is a statistically insignificant relationship between post CS wound sepsis and HIV among women this is confirmed by their p-values of 0.805 and 0.731 respectively which is less than the alpha value of 0.05. Therefore, we cannot reject the null hypothesis that states that there is no significant relationship between post CS wound sepsis and HIV among women and we reject the alternate hypothesis that there is a significant relationship. In summary therefore we can state that there is no evidence that HIV is a risk factors that may determine post CS wound sepsis among women. In summary therefore we can state that there is no evidence that HIV is a risk factors that may determine post CS wound sepsis among women.

**Table 4:** Diabetes as a risk factor for post-CS wound sepsis.

| Independent variable | Post-CS wound Sepsis | | $\chi^2$<br>(Pearson,<br>Likelihood ratio) | p-value |
| --- | --- | --- | --- | --- |
|  | yes | no |  |  |
| Diabetes |  |  | 0.386, 0.728 | 0.535, 0.394 |
| Yes | 0 | 6 |  |  |
| No | 6 | 93 |  |  |

From the result above the non-parametric Pearson chi-square statistic is 0.387 and the likelihood ratio statistics is 0.728 with a corresponding asymptotic p-value of 0.535 and 0.394 respectively. However, the result shows that there is a statistically insignificant relationship between post CS wound sepsis and diabetes among women this is confirmed by their p-values of 0.535 and 0.394 respectively which is more than the alpha value of 0.05. Therefore, we fail to reject the null hypothesis that states that there is no significant relationship between post CS wound sepsis and age distribution of women and we reject the alternate hypothesis that there is a significant relationship. summary therefore we can state that there is no evidence. In summary therefore we can state that there is no evidence that diabetes is a risk factors that may determine whether or not women may contract post CS wound sepsis.

**Table 5:** Hypertension as a risk factor for post-CS wound sepsis.

| Independent variable | Post-CS wound Sepsis | | $\chi^2$<br>(Pearson,<br>Likelihood ratio) | p-value |
| --- | --- | --- | --- | --- |
| Hypertension | yes | no | 0.001, 0.001 | 0.974, 0.974 |
| Yes | 1 | 5 |  |  |
| No | 16 | 83 |  |  |

From the result above the non-parametric Pearson chi-square statistic is 0.001 and the likelihood ratio statistics is 0.001 corresponding asymptotic p-values of 0.974. However, the result shows that there is a statistically insignificant relationship between post CS wound sepsis and hypertension among women this is confirmed by their p-values of 0.974 which is less than the alpha value of 0.05. Therefore, we fail to reject the null hypothesis that states that there is no significant relationship between post CS wound sepsis and hypertension among women and we reject the alternate hypothesis that there is a significant relationship. In summary therefore we can state that there is no evidence that hypertension is a risk factors that may determine post CS wound sepsis among women.

**Table 6:** Antepartum as a risk factor for post-CS wound sepsis.

| Independent variable | Post-CS wound Sepsis | | $\chi^2$<br>(Pearson,<br>Likelihood ratio) | p-value |
| --- | --- | --- | --- | --- |
| Antepartum infection | yes | no | 0.104, 0.100 | 0.747, 0.752 |
| Yes | 2 | 4 |  |  |
| No | 27 | 72 |  |  |

From the result above the non-parametric Pearson chi-square statistic is 0.104 and the likelihood ratio statistics is 0.100 corresponding asymptotic p-values of 0.747 and 0.752 respectively. However, the result shows that there is a statistically insignificant relationship between post CS wound sepsis and antepartum infection among women this is confirmed by their p-values of 0.747 and 0,752 respectively which is less than the alpha value of 0. 05. Therefore, we cannot reject the null hypothesis that states that there is no significant relationship between post CS wound sepsis and antepartum infection among women and we reject the alternate hypothesis that there is a significant relationship. In summary therefore we can state that there is no evidence that antepartum infection is a risk factors that may determine post CS wound sepsis among women.

**Table 7:** Established labour prior to CS as a risk factor for post-CS wound sepsis.

| Independent variable | Post-CS wound Sepsis | | $\chi^2$<br>(Pearson,<br>Likelihood ratio) | p-value |
| --- | --- | --- | --- | --- |
|  | yes | no |  |  |
| Established labour prior to CS |  |  | 0.044, 0.046 | 0.833, 0.830 |
| Yes | 1 | 5 |  |  |
| No | 20 | 79 |  |  |

From the result above the non-parametric Pearson chi-square statistic and the likelihood ratio statistics have a corresponding asymptotic p-values of 0.044 and 0.046 respectively. However, the result shows that there is a statistically insignificant relationship between post CS wound sepsis and Hb among women this is confirmed by their p-values of 0.833 and 0,830 respectively which is less than the alpha value of 0. 05. Therefore, we cannot reject the null hypothesis that states that there is no significant relationship between post CS wound sepsis and labour prior to CS among women and we reject the alternate hypothesis that there is a significant relationship. In summary therefore we can state that there is no evidence that labour prior to CS is a risk factors that may determine post CS wound sepsis among women.

**Table 8:** Established PROM prior to CS as a risk factor for post-CS wound sepsis.

| Independent variable | Post-CS wound Sepsis | | $\chi^2$<br>(Pearson,<br>Likelihood ratio) | p-value |
| --- | --- | --- | --- | --- |
|  | yes | no |  |  |
| Established PROM prior to labour |  |  | 1.023, 0.759 | 0.312, 0.383 |
| Yes | 1 | 5 |  |  |
| No | 6 | 93 |  |  |

From the result above the non-parametric Pearson chi-square statistic and the likelihood ratio statistics have a corresponding asymptotic p-values of 1.023 and 0.759 respectively. However, the result shows that there is a statistically insignificant relationship between post CS wound sepsis and patient reported outcome (prom) among women this is confirmed by their p-values of 0.312 and 0,383 respectively which is less than the alpha value of 0. 05. Therefore, we cannot reject the null hypothesis that states that there is no significant relationship between post CS wound sepsis and prom among women and we reject the alternate hypothesis that there is a significant relationship. In summary therefore we can state that there is no evidence that prom is a risk factors that may determine post CS wound sepsis among women.

## CHAPTER FIVE DISCUSSION

This study is a retrospective audit of all patient records of women who delivered by CS at Family Health Hospital (FHH) to ascertain the prevalence and factors associated with post-Caesarean Section wound sepsis at family health hospital, Teshie, greater Accra region.

Out of the 105 who delivered by CS during the 1-year study period, 6 women were identified to have post-CS wound sepsis identified translating a prevalence of post-CS wound sepsis to be 5.71%. This agrees with the current post CS prevalence rate of 3.7 to 24.2% (Wloch, C., et al., 2012).

However, studies in some Low-and-Middle-Income Countries (LMIC) such as India and Brazil reported incidence rates of 24.2% and 11-25% respectively (Cadrso, et al.,2010) which is higher than the results from this study.

Firstly, the differences in the prevalence rates reported in these studies can be attributed to the fact that this study had a retrospective design and identified women who presented to FHH with post-Caesarean Section wound sepsis prior to / after initial discharge, thus, the possibility exists that the index study was biased towards identifying women with more severe post CS sepsis. This could explain our lower incidence compared to other low-income countries.

Furthermore, the lower rates connote to adequacy and high quality of care among Caesarean births, strict adherence to infection protocols and aseptic technique, wound management and conditions in the theatres and the laying-in wards in FHH, given that the comparative study was conducted 8 years ago.

The highest participating age group was within 31 and 40 years (69.5%) (n=73). The median age of the patient population was 33 years while the mean age was 33.7 years.

### Specific risk factors for post CS sepsis

Despite including all the women who delivered by Caesarean section in the 1-year period in this study (114 in total), the number of women diagnosed with post CS sepsis was small – only 6 women. Due to this small number, identifying specific risk factors proved difficult, with all factors not reaching statistical significance. However, interesting trends towards significance were identified.

#### Age

The highest participating age group was within 31 and 40 years (69.5%) (n=73). The median age of the patient population was 33 years while the mean age was 33.7 years. Younger age and nulliparity have been positively correlated with increased risk of post CS wound sepsis even though the cause remains unclear (Wloch, C., et al., 2012). This explains why age was not found to be a significant risk factor for post-CS wound sepsis (p-value = 0.170) as majority of the study population was not of a younger age (< 30)

#### Anaemia

Anaemia has commonly been described as an associated factor for the development of post-operative sepsis. However, in the index study anaemia as risk factor did not reach statistical significance (p-value = 0.285). This is likely due to a very low prevalence of anaemia, given that only 3 (2.86%) patients with post-CS wound sepsis presented with a Hb below 11g/dL. This contrasts sharply with a South Africa research, where anaemia was present in 39.4% who later succumbed to pregnancy-related wound sepsis. When severe, anaemia can cause reduced resistance to sepsis and slower recovery from infection. (Olsen, et al., 2010).

#### HIV

HIV infection was not found to have any statistical association in this study (p=0.805). This study had no woman who had HIV infection and post-wound CS sepsis. Women living with HIV infection have higher risk of post CS wound sepsis especially when they are not yet on HAART or have low CD4 count, WHO stage 3 or 4 disease or with opportunistic infections (Coetzer, M., 2017). It is possible that the only woman who was found to be HIV was on HAART and as such wasn’t vulnerable to opportunistic infections that will result in post-CS wound sepsis.

#### Diabetes mellitus

Diabetes was not found to be a statistically significant risk factor for post CS sepsis (p-value = 0.535). This is likely due to the fact that no woman was identified with diabetes and subsequently being diagnosed with post-wound CS sepsis. Schneid-Kofman et al. found that the highest risk of sepsis exists in women with pre-gestational diabetes, but that women with gestational diabetes are also at increased risk. The study group acknowledges this proven increased risk, but postulates that diabetes in pregnancy, in our study population, is not one of the major contributors to the burden of post CS sepsis. Great care is taken in the FHH setting to achieve optimal glycaemic control, in order to decrease other risks related to diabetes in pregnancy. The great majority of women have well-controlled diabetes with little, or no target-organ involvement by the time of delivery. This, together with the relatively low numbers in general, might explain the lack of statistical significance in this study.

#### Hypertension

Hypertension was found not to be a statistically significant risk factor for post CS sepsis (p-value = 0.974). This can be attributed to a very low number (1) of hypertensive disorders with post-CS wound sepsis in the study population. Women suffering from hypertensive disorders in pregnancy have been shown to have an increased risk of developing post CS wound sepsis (Olsen, et al., 2010). This can be explained by the great care taken in the FHH setting to achieve blood pressure control, in order to decrease other risks related to hypertension in pregnancy.

#### Antepartum infection

The presence of active antepartum infection, such as chorioamnionitis, urinary tract infections has been well established as a significant independent risk factor for post CS sepsis, especially endometritis (Alan T.N.T. & Andrews W.W. (2010). However, the index study did not yield a statistically significant (p-value = 0.747) result as only 6 women presented with antepartum infection and of these, only 2 women also had evidence of sepsis after CS. This could indicate that antenatal infection is not the most significant contributor to sepsis in the FHH setting and that women in this study population rather develop sepsis due to multiple other risk factors, however numbers are too small to draw accurate conclusions.

#### Active labour prior to CS

A trend towards developing post CS sepsis was identified in women with longer durations of active labour. Ghuman et al. found the presence of active labour prior to CS to be an independent risk factor for post CS sepsis. The study group acknowledges the previously proven association between active labour and post CS sepsis and postulates that this was not replicated in the index study for two reasons. Firstly, due to very few women with both active labour and sepsis included (only 1 out of 114 women), it is very difficult to draw accurate statistical conclusions. Secondly, there could be other conditions that will make a pregnancy high-risk, thus, a woman without active labour might have numerous other risk factors and therefore develops sepsis due to other factors.

#### Established PROM prior to labour

The presence of active antepartum infection, such as chorioamnionitis, urinary tract infections has been well established as a significant independent risk factor for post CS sepsis, especially endometritis (Alan T.N.T. & Andrews W.W. (2010). However, the index study did not yield a statistically significant (p-value = 0.747) result as only 6 women presented with antepartum infection and of these, only 2 women also had evidence of sepsis after CS. This could indicate that antenatal infection is not the most significant contributor to sepsis in the FHH setting and that women in this study population rather develop sepsis due to multiple other risk factors, however numbers are too small to draw accurate conclusions.

### 5.1 STRENGTH AND LIMITATIONS

This study was the first audit done on post CS sepsis in the FHH setting. It included a relatively large number of women and was well powered to accurately estimate the post CS sepsis rate.

Firstly, due to the retrospective nature of the study design, it relied heavily on accurate note keeping and a number of patients had to be excluded due to inadequate or unavailable notes. In addition, some valuable information wasn’t included in the notes as such it made it difficult to determine the risk associated with such factors.

Despite including all women that delivered within the study period, the results yielded a lower than expected post CS sepsis rate, although this can be interpreted as a reassuring finding – it was still difficult to determine statistically significant risk factors specific to this population accurately due to the low number of post-CS sepsis.

The study group initially expected to find clear associations between known risk factors (raised BMI, hypertension, diabetes, HIV, etc.) and post CS sepsis, however, this was not the case. This raises awareness that due to the patient profile and multifactorial aetiology, even women that are usually regarded as “lower risk” (for example non-HIV infected women, women with a normal BMI, non-diabetes, non-hypertensives, etc) are at significant risk of post CS sepsis.

### 5.2 CONCLUSION

Despite a post-CS sepsis prevalence that compares well with the current global prevalence (35.7% vs. 3.7 – 24.2%), post-CS sepsis remains a significant contributor to maternal morbidity in the Ghanaian setting. Risk factors for post-CS sepsis remain multifactorial and in the setting of a Family Health Hospital (FHH), a private hospital located in Teshie - all women should be treated as potentially at risk. Optimization of chronic medical conditions, vigilant intra-partum care, meticulous surgical technique and recognition of early signs of post-CS sepsis are essential in order to prevent maternal morbidity. Further prospective research is needed in order to identify specific, individual, modifiable risk factors.

### 5.3 RECOMMENDATION

A prospective, matched case-control study is advised in order to assess whether trends in this study reach a statistical significance when adequately powered. Qualitative research into patient perception of post CS sepsis, the impact on quality of life as well as barriers to seeking early medical intervention should be undertaken.

Also, clinicians should be educated on the need to make comprehensive notes in order to aid other retrospective studies as such studies relies completely on this secondary data to achieve its objectives.

Furthermore, the hospital software providers may need to update their software from time-time as this may be one of the reasons for lack of comprehensive notes as it was cumbersome to work with.

## Supporting information

IRB Approval

## Data Availability

All data produced in the present work are contained in the manuscript

## REFERENCES

Alan, T.N.T., & Andrews, W.W. (2010). Diagnosis and Management of Clinical Chorioamnionitis. Clinical Perinatology, 37(2): 339–354. 10.1016/j.clp.2010.02.003

Alanis, M. C., Villers, M. S., Law, T. L., Steadman, E. M., & Robinson, C. J. (2010). Complications of cesarean delivery in the massively obese parturient. American journal of obstetrics and gynecology, 203(3), 271.e1–271.e2717. 10.1016/j.ajog.2010.06.049

Betran, A.P., Jianfeng, Ye., Moller, A-B., Zhang, J., Guelmezoglu, A.M., Torloni, M.R. (2016). The Increasing Trend in Caesarean Section Rates: Global, Regional and National Estimates: 1990-2014. PLOS ONE 11(2): e0148343. 10.1371/journal.pone.0148343

Bhavani, K., Prasanthi, S., Jyothsna, Y., Vani, I., Uma, N. (2017). A critical review of post-operative caesarean section sepsis - A retrospective study. IAIM, 4(11): 153–159.

Cardoso, M.C., Pinto, N.A.M. (2010). Post-discharge surveillance following caesarean section: the incidence of surgical site infection and associated factors. American Journal Infection Control, 38(6):46772. Retrieved from: http://www.ncbi.nlm.nih.gov/pubmed/2022657. Accessed on 6th August,2020.

Centre of Disease Control and Prevention, (2013). Centre Disease Control / National Healthcare Safety Network Protocol Clarifications. Retrieved from: www.cdc.gov accessed on 6th August,2020.

Chunder, A., Devjee, J., Khedun, S.M., Moodley, J., Esterhuizen, T. (2012). A randomised controlled trial of suture materials used for caesarean section skin closure: Do wound infection rates differ? South African Medical Journal. 2012;102(6):374–6.

Chu, K., et al., (2012). Cesarean section rates and indications in sub-Saharan Africa: a multi-country study from Medecins sans Frontieres. PLOS one, 7(9).

Coetzer, M., (2017). A Retrospective Audit of Post-Caesarean Sepsis at Tygerberg Hospital. Accessed from https://scholar.sun.ac.za/bitstream/handle/10019.1/102951/coetzer_retrospective_2017.pdf?sequence=1&isAllowed=y on 8th September,2020.

Costa, T., Medeiros, P., Selles, M. (2018). Smoking increases the risk of surgical site infection after hydrocelectomy in adults: A retrospective cohort study in Brazil. Avaliable from : https://www.researchgate.net/publication/322379165_Smoking_increases_the_risk_of_surgical_site_infection_after_hydrocelectomy_in_adults_A_retrospective_cohort_study_in_Brazil accessed on 8th September 2020.

Devjani, D., Sonal, S., Geeta, M., Yadav, R., Dutta, R. (2013). Risk Factor Analysis and Microbial Etiology of Surgical Site Infections following Lower Segment Caesarean Section. International Journal Antibiotics Agents (Art ID: 283025) :1–6. Available from: http://www.hindawi.com/journals/ijan/2013/283025/. Accessed on 6th August, 2020.

Dillen, J., Zwart, J., Schutte, J.M., Roosmalen, J. (2012). Maternal Sepsi: Epidemiology, Etiology and Outcome. mhttps://www.researchgate.net/publication/43073830_Maternal_Sepsis_Epidemiology_Etiology_And_Outcome Accessed on 8th September,2020.

Dlamini, L.D., et al., (2015). Antibiotic prophylaxis for caesarean section at a Ugandan hospital: a randomised clinical trial evaluating the effect of administration time on the incidence of postoperative infections. BMC pregnancy and childbirth. 5(1): p. 91.

Ezechi, O.C., Edet, A., Akinlade, H., Gab-Okafor, C.V., Herbertson, E. (2009). Incidence and risk factors for caesarean wound infection in Lagos Nigeria. BMC Research Notes. 2:186.

Gelaw, K.A., et al., (2017). Surgical site infection and its associated factors following caesarean section: a cross sectional study from a public hospital in Ethiopia. Patient safety in surgery, 11(1): p. 18.

Ghana Demographich Health Survey. (2014). Available from: https://dhsprogram.com/pubs/pdf/FR307/FR307.pdf Accessed on 8th September 2020.

Ghuman, M., Rohlandt, D., Joshy, G., Lawrenson, R. (2011). Post-caesarean section surgical site infection: rate and risk factors. New Zealand Medical Journal., 124(1339):32–6.

Goldenberg, R.L., & Harrison, M.S. (2016). Caesarean section in sub-Saharan Africa. Maternal health, neonatology and perinatology, 2(1): p. 6.

Hafeez, M., Yasin, A., Badar, N., Pasha, M.I, Akram, N., Gulzar, B. (2014). Prevalence and indication of caesarean section in a teaching hospital. JIMSA., 27(1):15–6.

Irani, M., & Deering, S. (2015). Challenges affecting access to caesarean delivery and strategies to overcome them in low-income countries. International Journal of Gynaecology & Obstetrics, 131(1): p. 30–34.

Kateete, D.P., et al., (2011). High prevalence of methicillin resistant Staphylococcus aureus in the surgical units of Mulago hospital in Kampala, Uganda. BMC research notes, 4(1): p. 326.

Krieger, Y., Walfisch, A., Sheiner, E. (2017). Surgical site infection following cesarean deliveries: trends and risk factors. The journal of maternal-fetal & neonatal medicine: the official journal of the European Association of Perinatal Medicine, the Federation of Asia and Oceania Perinatal Societies, the International Society of Perinatal Obstetricians, 30(1), 8–12. 10.3109/14767058.2016.1163540

Leth, R.A., Uldbjerg, N., Nørgaard, M., Møller, J.K., Thomsen, R.W. (2011). Obesity, diabetes, and the risk of infections diagnosed in hospital and post-discharge infections after caesarean section: a prospective cohort study. Acta Obstetricia Gynecologica Scandinavica, 90(5):501–9. Available from: http://www.ncbi.nlm.nih.gov/pubmed/21306347. Accessed on 6th August, 2020.

Manyeh, A.K., Amu, A., Akpakli, D.E., Williams J., Gyapong M., (2018). Caesarean section delivery in Southern Ghana: evidence from INDEPTH Network project. ORCID: orcid.org/0000-0002-7005-57071,2. BMC Pregnancy and Childbirth volume 18.

Mitchell M.D., et al., (2015). Placental exosomes in normal and complicated pregnancy. American journal of obstetrics and gynaecology, 213(4): p. S173–S181.

Muhumuza, I., Abdulrahman, Z.L., Tayebwa, B., et al. (202). Post Caesarean Wound sepsis and associated factors among patients attending a rural regional referral hospital in Western Uganda: A cross-sectional study, available at Research Square 10.21203/rs.2.24374/v1 accessed on 8th September, 2020

Nimbalkar, A. H. (2015). A study to compare the efficacy of subcutaneous suture re-approximation versus subcutaneous drain for the prevention of wound complication in women undergoing caesarean section. Journal of Evidence page medicine and health care, 2(43):7789–7797 10.18410/jebmh/2015/1050

Olsen, M.A., Butler, A., Willers, D., Gross, G.A., Devkota, P., Fraser V. (2010). Risk factors for endometritis following low transverse caesarean section. Infect Control Hosp Epidemiology, 31pg69–77.

Opøien, H.K., Valbø, A., Grinde-Andersen, A., Walberg M. (2007). Post-caesarean surgical site infections according to CDC standards: rates and risk factors. A Prospective Cohort Study. Acta Obstetricia Gynecologica Scandinavica, 86 (9): 1097 – 102. Available from: http://www.ncbi.nlm.nih.gov/pubmed/17712651. Accessed on 6th August, 2020.

Pevzner, L., Swank, M., Krepel, C., Wing, D. A., Chan, K., Edmiston CE. (2011). Effects of maternal obesity on tissue concentrations of prophylactic cefazolin during cesarean delivery. Obstetrics and Gynaecology 117(4): 877 – 882.10.1097/AOG.0b013e31820b95e4

Schneid-Kofman, N., Sheiner, E., Holcberg, L. G. (2005). Risk factors for wound infection following cesarean deliveries. International Journal of Obstetrics and Gynecology., 90(1):10–5. Available from: http://www.ncbi.nlm.nih.gov/pubmed/15913620. Accessed on 6th August, 2020.

Skele, E., Koch A.M., Harthug, S., Fosse, U., Sygnestveit, K., Nilsen, R.M., Tangvik, R.J. (2018). A positive association between nutritional risk and the incidence of surgical site infections: A hospital-based register study. 10.1371/journal.pone.0197344

Tsai, P.-S., Hsu, C.-S., Fan, Y.-C., Huang, C.-J. (2011). General anaesthesia is associated with increased risk of surgical site infection after Caesarean delivery compared with neuraxial anaesthesia: a population-based study. British Journal of anaesthesia, 107(5):757–61. 10.1093/bja/aer262

Tuuli, M.G., Liu, J., Stout, M.J, Martin, S., Cahill, A.G., Odibo, A.O., et al., (2016). A Randomized Trial Comparing Skin Antiseptic Agents at Cesarean Delivery. New England Jounal of Medicine, 374(7):647–55. Available from: http://www.ncbi.nlm.nih.gov/pubmed/26844840. Accessed on 6th August, 2020.

Wloch, C., Wilson, J.A., Lamagni, T., Harrinton, P., (2012). Risk factors for surgical site infection following caesarean section in England: Results from a multicentre cohort study. Available from https://www.researchgate.net/publication/230614674_Risk_factors_for_surgical_site_infection_following_caesarean_section_in_England_Results_from_a_multicentre_cohort_study accessed on 8th Spetember,2020

Ziogos, E., Tsiodras, S., Matalliotakis, I., Giamarellou, H., & Kanellakopoulou, K. (2010). Ampicillin/sulbactam versus cefuroxime as antimicrobial prophylaxis for cesarean delivery: a randomized study. BMC infectious diseases, 10, 341. 10.1186/1471-2334-10-341

Zuarez-Easton, S., et al., (2017). Post caesarean wound infection: prevalence, impact, prevention, and management challenges. International journal of women’s health, 9: p. 81.

