## Supplementary material for "Prevalence And Factors Associated with Post-Caeserean Section Wound Sepsis in A Hospital in Ghana (A Retrospective Audit)": IRB Approval

UNIVERSITY OF GHANA
MEDICAL SCHOOL
DEPARTMENT OF COMMUNITY HEALTH

September 14, 2020

**Student Name:** **Wendy Muonibeh Bebobru**
**Proposal Id:** **UGMS-CHDRC/013/2020**
**Title:** **Prevalence and Factors Associated with Post-Casearean Section Wound Sepsis at Family Health Hospital, Teshie, Greater Accra Region. (A Retrospective Audit)**

**PROPOSAL AND ETHICAL APPROVAL**

The Community Health Department Review Committee (CHDRC) of the University of Ghana Medical School, has reviewed and given approval for the conduct of the above research by the final year Medical Student.

Progress of the research will be monitored and final output will be reviewed and assessed in partial fulfillment of the Final MBCh.B Examination of the Medical School.

Thank you.

Yours sincerely,

For

Prof. Alfred E. Yawson
(The Chairman / Head of Department, Community Health)
